# Digital Epidemiology for Epidemic Forecasting: Evaluating Twitter Emotion Signals and Google Symptom Searches Using Prophet and SARIMAX Models

**DOI:** 10.64898/2026.01.18.26344365

**Authors:** Thierry Guigma, Ian Brooks

## Abstract

**Background:** Digital traces from social media and online search platforms have been used to support infectious disease forecasting, but their performance during the COVID-19 pandemic has varied widely. It is still uncertain which types of digital signals add dependable information to established forecasting models.

**Objective:** This study evaluated whether Twitter indicators and Google symptom search trends improve forecasts of COVID-19 cases in the United States. A second goal was to examine whether any gains remain consistent across two different forecasting approaches, Prophet and SARIMAX.

**Methods:** National daily COVID-19 case data were linked with Twitter-based emotion and Google Trends symptom variables. A forward-selection procedure identified the strongest predictors from each source. Four Prophet models were trained and tested through rolling 30-day forecasts. The same predictor sets were then used in parallel SARIMAX models. Performance was assessed using RMSE, MAE, and MAPE, and results were inspected across major epidemic waves.

**Results:** Twitter indicators produced the clearest and most consistent improvements. In the Prophet models, the Twitter-enhanced version reduced 30-day forecast error by about 14% compared with the baseline. Google symptom searches showed smaller and less stable improvements, and combining Google trends with Twitter signals did not produce additional benefits. SARIMAX models showed the same general pattern, although improvements were more modest. Across epidemic waves, Twitter-based models reacted more quickly to shifts in transmission than the baseline model.

**Conclusions:** Twitter emotion indicators, especially neutral and informational posts, provided meaningful forecasting value across models and horizons. Google symptom searches contributed far less and did not strengthen performance when added to the Twitter predictors. The consistency of the findings across two modeling frameworks suggests that social media activity can offer reliable supplemental information for real-time epidemic forecasting. Continued work is needed to understand how these signals behave at finer spatial scales and in future outbreaks.

## INTRODUCTION

Digital epidemiology has rapidly expanded over the past decade (1, 2). Social media activity, search engine queries, mobility traces, and other digital signals can provide real-time insights into population behavior and early signs of emerging outbreaks (3–5). During the COVID-19 pandemic, these sources became especially valuable as traditional surveillance systems experienced delays, under-reporting, and disruptions in testing capacity (6). As a result, multiple research groups investigated the feasibility of using digital traces to supplement conventional forecasting and improve situational awareness (7–9).

Among digital data sources, X (will be referred as Twitter) and Google Trends have been the two most frequently studied platforms. Twitter provides timely, high-volume public discourse reflecting population concern, symptom reporting, emotional responses, and behavioral intentions (10–13). Prior work has shown that sentiment and emotion signals extracted from tweets can serve as early indicators of changes in epidemic trajectories and public risk perception (14–16). Google Trends, in contrast, captures collective information-seeking behavior and symptom searches, which may precede clinical presentation or testing (17–19). Early in the pandemic, studies demonstrated that searches for symptoms such as “loss of smell,” “fever,” or “COVID test” correlated with (and anticipated in some settings) changes in confirmed cases or hospitalization trends (20–22).

Despite growing interest, evidence on the predictive value of these digital signals remains mixed. Several studies reported strong correlations between digital indicators and COVID-19 outcomes, but weaker performance when used directly as forecasting regressors (15, 23–25). Google Trends-based models have been particularly inconsistent across countries, time periods, and symptom keywords (26–28). Twitter-based forecasting has shown promise, especially when using emotion or sentiment features, but few studies have compared Twitter and Google signals directly or evaluated their incremental contribution within the same modeling framework (11, 14, 29). Moreover, most digital epidemiology studies rely on a single forecasting model, making it difficult to determine whether observed effects are model-specific or robust across methodological approaches.

At the same time, COVID-19 remains a relevant case study for evaluating digital surveillance methods. Although vaccination coverage has increased and global case counts have declined, the virus continues to circulate, and new variants periodically emerge. COVID-19 forecasting remains an important benchmark for testing digital epidemiology methods given its long time series, multiple epidemic waves, and abundant digital data availability. Insights from COVID-19 forecasting research can also extend to future respiratory pathogens and emerging infectious diseases.

This study provides a comprehensive evaluation of Twitter emotion signals and Google symptom searches for COVID-19 forecasting in the United States. We compare these digital indicators within two distinct modeling approaches: Prophet, a nonlinear additive model widely used for time-series forecasting, and SARIMAX, a classical autoregressive model with exogenous regressors. By applying the same sets of regressors across both model families and evaluating performance using rolling-origin cross-validation, we assess whether the predictive value of digital signals is consistent across models. We also looked at how forecast performance varied across major epidemic waves and how accuracy changed with the length of the prediction horizon.

Our contribution lies in (1) directly comparing Twitter and Google signals within the same methodological pipeline, (2) validating results across two fundamentally different model classes, and (3) providing wave-specific analyses that reveal when digital indicators are most informative. This work advances the understanding of digital epidemiology signals and their potential role in enhancing real-time infectious disease forecasting.

## METHODS

### Study Design

We constructed parallel forecasting frameworks using both the Prophet algorithm and Seasonal Autoregressive Integrated Moving Average with Exogenous Regressors (SARIMAX). This dual-model strategy was chosen to evaluate whether the predictive contribution of digital signals was consistent across additive trend-seasonality models and classical statistical time-series approaches, an issue highlighted in prior digital epidemiology literature (3–8). Prophet and SARIMAX differ fundamentally in how they represent trend, seasonality, and temporal dependence. Prophet models the time series as an additive combination of flexible, piecewise-linear (or logistic) trends, multiple seasonal components, and optional holiday effects, allowing structural changes in trend and seasonality to be learned directly from the data with minimal stationarity assumptions (30, 31). In contrast, SARIMAX is a parametric autoregressive framework that assumes a stationary or differenced process, with seasonality captured through fixed seasonal autoregressive and moving-average terms and temporal dependence explicitly encoded via lag structures (32, 33). As a result, Prophet is better suited to capturing abrupt trend changes and nonlinear epidemic dynamics, whereas SARIMAX emphasizes short-term autocorrelation and persistence in the observed series (34, 35). Evaluating digital indicators across both frameworks therefore helps determine whether their predictive contribution reflects robust external information or depends on how trend and seasonality are modeled. Using two contrasting approaches also provides a more reliable assessment of how stable these signals are when applied to real-world forecasting tasks.

### Data Sources

#### COVID-19 Case Data

Daily confirmed COVID-19 cases for the United States were retrieved from the U.S. Centers for Disease Control and Prevention (CDC) COVID Data Tracker (36, 37). This dataset offers laboratory-confirmed case counts aggregated at the national level.

#### Twitter Digital Emotion Indicators

Twitter-based digital signals were obtained from the Open ICPSR COVID-19 Twitter Repository (38), which aggregates approximately 54 million pandemic-related tweets originating from the US. The repository provides daily counts of emotions (No Emotion, Fear, Anger, Sadness, Happiness) and sentiments (Very Negative, Negative, Neutral, Positive, Very Positive) derived using supervised machine-learning models applied to tweet text.

All tweets were aggregated at daily resolution to align with the national COVID-19 case time series.

#### Google Symptom Search Trends

Symptom-related search data were extracted from the Google COVID-19 Symptoms Search Dataset, a curated repository derived from Google Trends (39, 40). This dataset includes normalized daily search intensities in various languages for clinically relevant symptoms such as cough, fever, sore throat, shortness of breath and anosmia, indicators that have demonstrated plausible early associations with COVID-19 incidence and hospitalization trends in prior studies (21, 25, 41). Only US-based searches were considered.

COVID-19 case reporting, Twitter activity, and Google search behavior all evolved over the course of the pandemic. These shifts introduce noise that may affect how strongly each signal relates to actual transmission. The analysis therefore relies on rolling cross-validation to evaluate performance across multiple phases of the epidemic rather than depending on any single period.

### Selection of External Regressors

To identify the most informative digital predictors, we applied a forward-selection procedure based on out-of-sample forecast accuracy. Candidate variables from each digital source were added incrementally to the baseline model and evaluated using rolling-origin forecasts. At each step, the predictor that produced the largest reduction in root mean squared error (RMSE) was retained. The selection process continued until no remaining candidate produced further improvements in RMSE. This stopping rule favors parsimonious models and limits overfitting, particularly when working with correlated and noisy digital indicators. The forward-selection routine was used to avoid overly complex models and to ensure that each added predictor provided a measurable contribution.

### Prophet Forecasting Models

Four Prophet models were estimated:

(1) a historical baseline using case data only;
(2) a Twitter-enhanced model;
(3) a Google-enhanced model; and
(4) a combined digital-signals model.

Cross-validated forecasts were generated using rolling-origin evaluation: 30-day forecasts repeated every 30 days to obtain a distribution of forecast errors. The set of external regressors included in each model was determined exclusively by the RMSE-based forward-selection procedure described above.

### SARIMAX Forecasting Models

To complement the Prophet models, we also built a set of SARIMAX models using the same sets of external regressors. The goal was to test whether the contribution of Twitter and Google indicators remained consistent across a more traditional time-series framework. We obtained a seasonal ARIMA model with weekly seasonality, reflecting the well-known 7-day reporting cycle in COVID-19 case data. This structure was kept the same for the baseline SARIMAX model and for all versions with added regressors, ensuring that any change in forecast performance could be attributed to the digital variables rather than to differences in model form.

### Performance Evaluation

Model accuracy was assessed using root mean squared error (RMSE), mean absolute error (MAE), and mean absolute percentage error (MAPE). For Prophet models, we examined how forecast error changed with prediction horizon by calculating RMSE at multiple lead times (e.g., 1 to 30 days). SARIMAX models were evaluated at the standard 30-day horizon.

### Reproducibility and Code Availability

All analyses were conducted in Python 3.10 using Jupyter Notebook. Forecasting models used the Prophet library (Meta; v1.1.5) and Statsmodels (v0.14) for SARIMAX. Data manipulation and visualization were performed using pandas (v2.1), numpy (v1.26), and matplotlib (v3.8). Hyperparameters, cross-validation settings, and modeling procedures followed fully reproducible code executed in a consistent environment.

All datasets used in this study are publicly available: the COVID-19 case data is from the U.S. Centers for Disease Control and Prevention (CDC), Twitter emotion variables from the Open ICPSR annotated COVID-19 Twitter dataset, and Google Trends symptom indices are from the COVID-19 Google Symptoms Search Dataset.

All analysis code, including preprocessing scripts, Prophet and SARIMAX models, and figure-generation routines, will be deposited in a public GitHub upon acceptance.

### Ethics Approval

This study did not involve human participants and used only publicly available, aggregate, de-identified datasets from CDC, Google, and publicly archived Twitter corpora. No individual-level health information, identifiable personal data, or private social media content were accessed. In accordance with U.S. federal regulations (45 CFR §46) and institutional policies, research using publicly available, de-identified datasets does not constitute human subjects research and therefore does not require Institutional Review Board (IRB) approval.

No interventions, interactions, or collection of personal data occurred during this study. All analyses complied with the data use policies of CDC, Google, and the Open ICPSR repository.

## RESULTS

### Descriptive Trends

From January 2020, the United States experienced several distinct COVID-19 waves, including the early 2020 surge, the winter 2020 to 2021 wave, the Delta wave of mid-2021, and the Omicron wave beginning in late 2021. Daily confirmed cases displayed pronounced nonlinearity, with abrupt accelerations and steep peaks during major outbreaks. Figure 1 presents the observed case trajectory across the study period.

**Figure 1:**
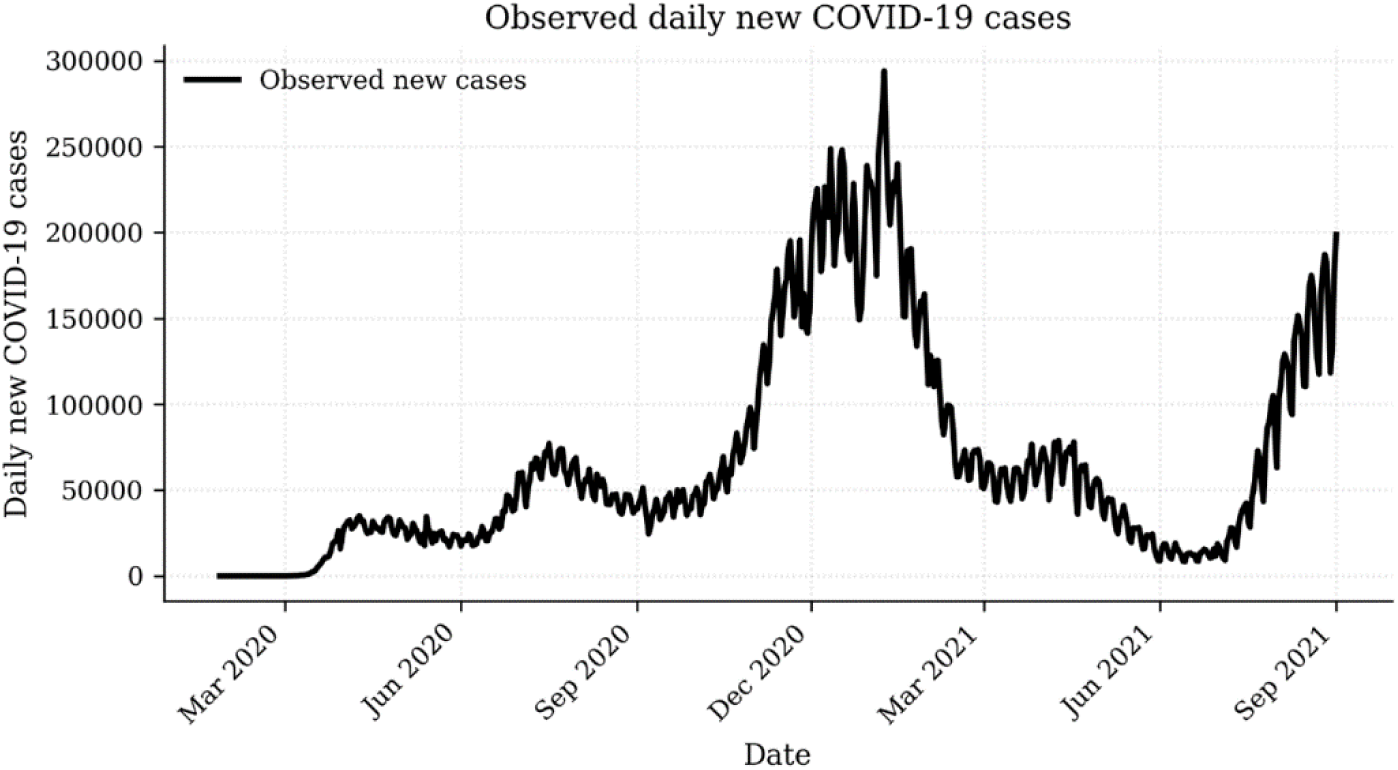
Observed daily new COVID-19 cases

### Prophet Model Performance

#### Forward-Selection of External Regressors

Forward selection identified three Twitter features as the strongest predictors: Neutral, VeryPositive, and NoEmotion. These variables produced the largest incremental reductions in RMSE when added to the Prophet model. For Google Trends, two symptoms, cough and anosmia, were identified as the best-performing regressors. Detailed forward-selection results are provided in Tables 1 and 2.

**Table 1:**
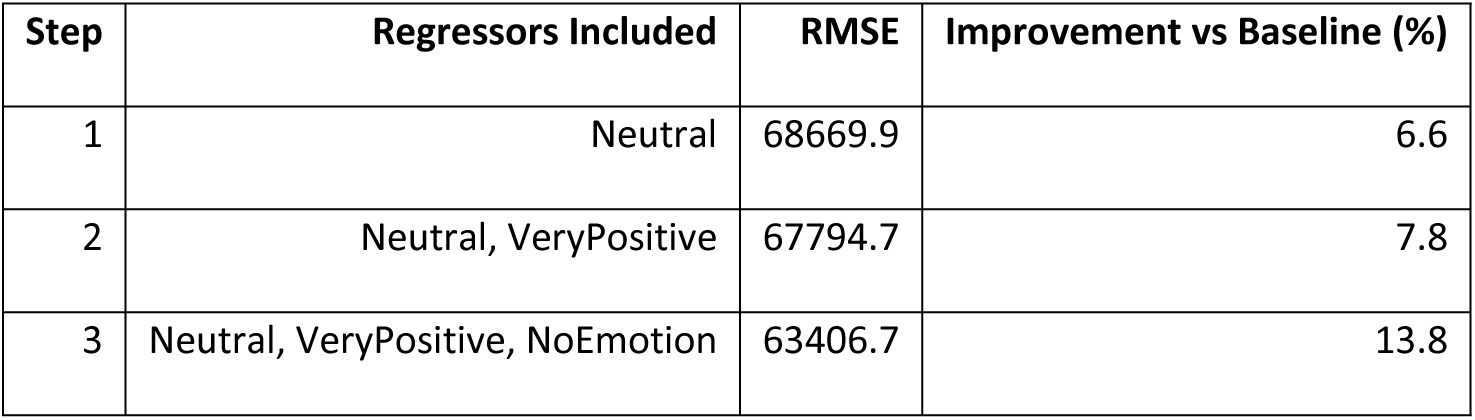
Forward-selection table for Twitter predictors.

**Table 2:**
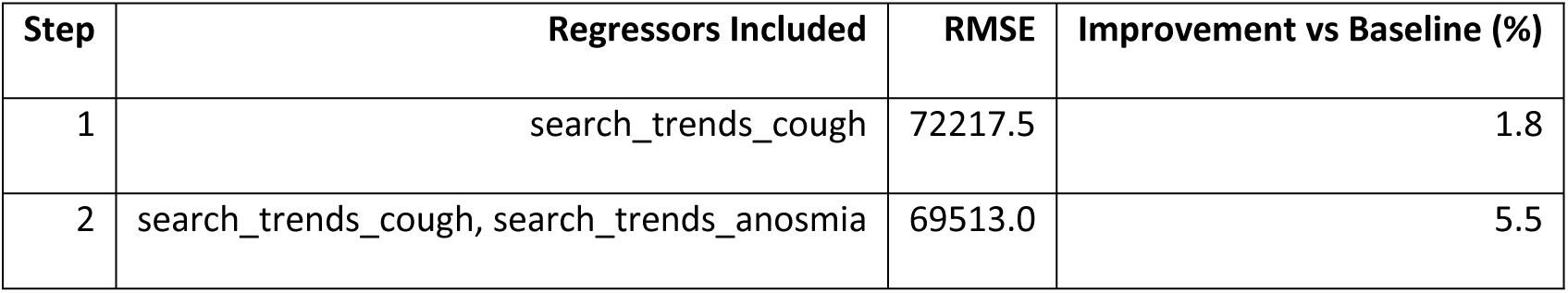
Forward-selection table for Google predictors.

| Step | Regressors Included | RMSE | Improvement vs Baseline (%) |
| --- | --- | --- | --- |
| 1 | search_trends_cough | 72217.5 | 1.8 |
| 2 | search_trends_cough, search_trends_anosmia | 69513.0 | 5.5 |

Although additional Twitter emotion and sentiment variables were evaluated during the forward-selection process, no further predictors reduced out-of-sample RMSE once Neutral, VeryPositive, and NoEmotion were included. Many Twitter variables are strongly correlated, and after the most informative signals were incorporated, remaining variables provided redundant information rather than incremental predictive value. As a result, adding further predictors did not improve forecast accuracy and was therefore avoided.

A similar pattern was observed for Google Trends variables. Only searches related to cough and anosmia improved RMSE, while other symptom searches failed to provide additional predictive gains once these variables were included.

### 30-Day Forecasting Performance

#### Prophet Baseline vs. Advanced Models

Performance for the four Prophet models is presented in Table 3.

**Table 3:** Model performance for 30-day rolling forecasts.

| Model | RMSE | MAE | MAPE | RMSE improvement (%) | MAE improvement (%) |
| --- | --- | --- | --- | --- | --- |
| Baseline (history only) | 73,555.0 | 47,640.5 | 0.50 | 0.0 | 0.0 |
| Twitter-enhanced Model | **63,406.7** | **40,750.6** | 0.40 | **13.8** | **14.5** |
| Google-enhanced Model | 69,513.0 | 43,332.2 | 0.40 | 5.5 | 9.0 |
| Twitter + Google Model | 69,259.9 | 43,367.6 | 0.40 | 5.8 | 9.0 |

**Table 4:** SARIMAX models’ performances.

| Model | RMSE | MAE | MAPE | RMSE improvement<br>(%) | MAE improvement<br>(%) |
| --- | --- | --- | --- | --- | --- |
| **SARIMAX Baseline** | 57,467.6 | 35,494.6 | 0.40 | 0.0 | 0.0 |
| **SARIMAX + Twitter** | **55,716.7** | **34,368.6** | 0.40 | **3.0** | **3.2** |
| **SARIMAX + Google** | 59,029.6 | 36,440.9 | 0.40 | -2.7 | -2.7 |
| **SARIMAX + Twitter +**<br>**Google** | 56,133.9 | 34,494.8 | 0.40 | 2.3 | 2.8 |

The baseline Prophet model produced an RMSE of approximately 73,555 and an MAE of roughly 47,640. Incorporating Twitter emotion indicators resulted in substantial improvements. The Twitter-enhanced model achieved an RMSE of about 63,407 and an MAE near 40,751. This represents a reduction in error of approximately 14% compared with the baseline. Google Trends regressors produced smaller and less consistent improvements. The Google-enhanced model lowered RMSE only to about 69,513 and produced modest reductions in MAE. The combined model that included both Twitter and Google regressors performed similarly to the Google-only model but with limited additional gains.

Figures 2 through 5 compare observed cases with forecasts from each Prophet model in full and zoomed views.

**Figure 2:**
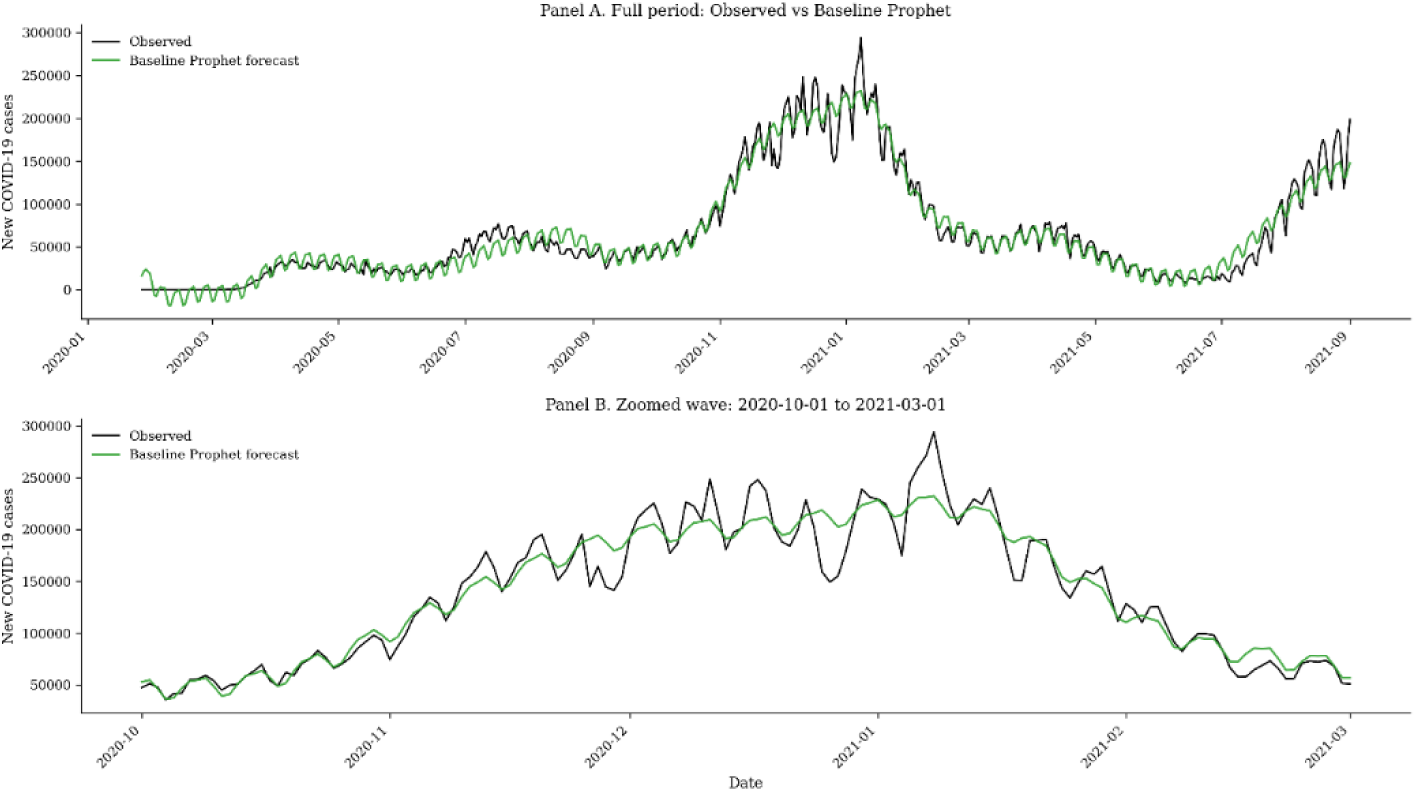
Observed vs Baseline forecast

**Figure 3:**
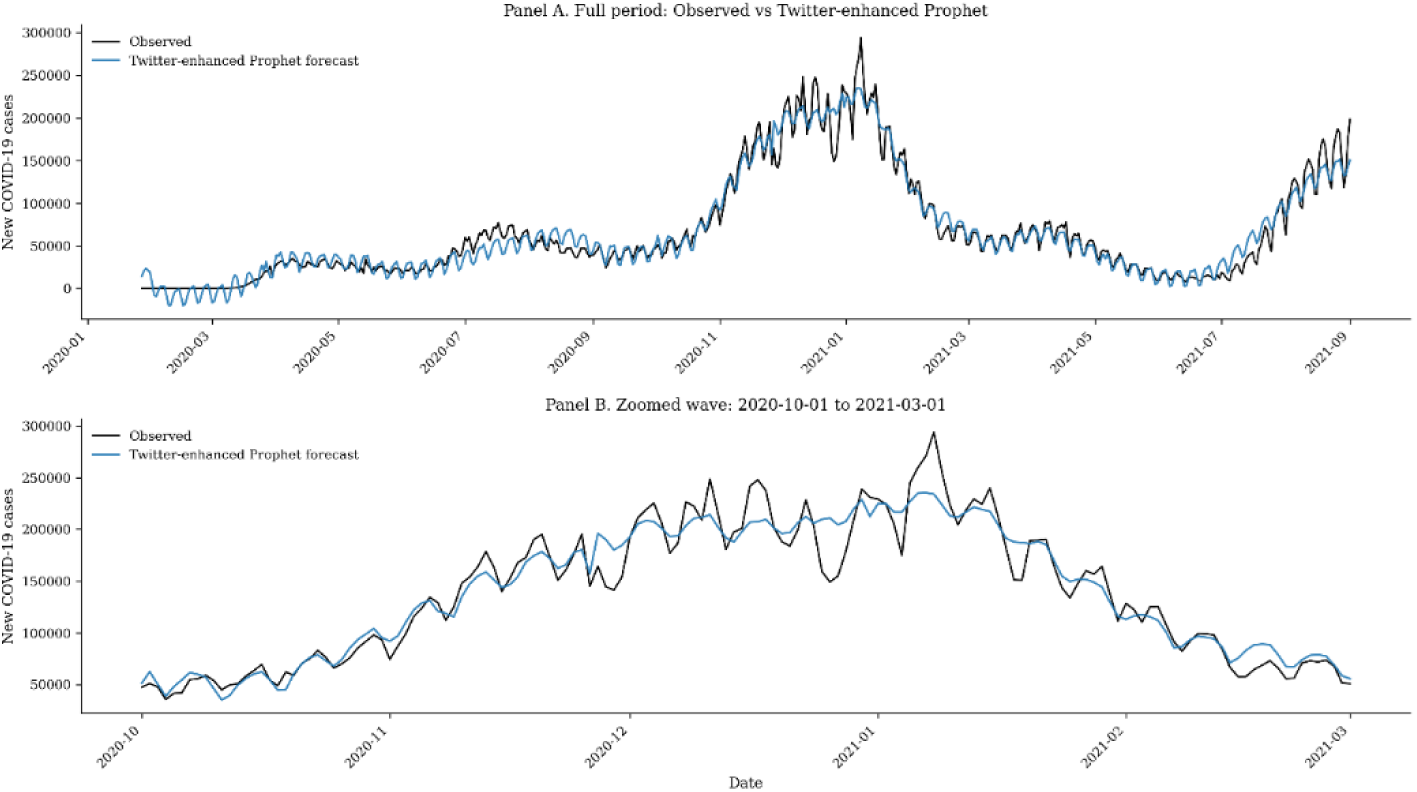
Observed vs Twitter forecast

**Figure 4:**
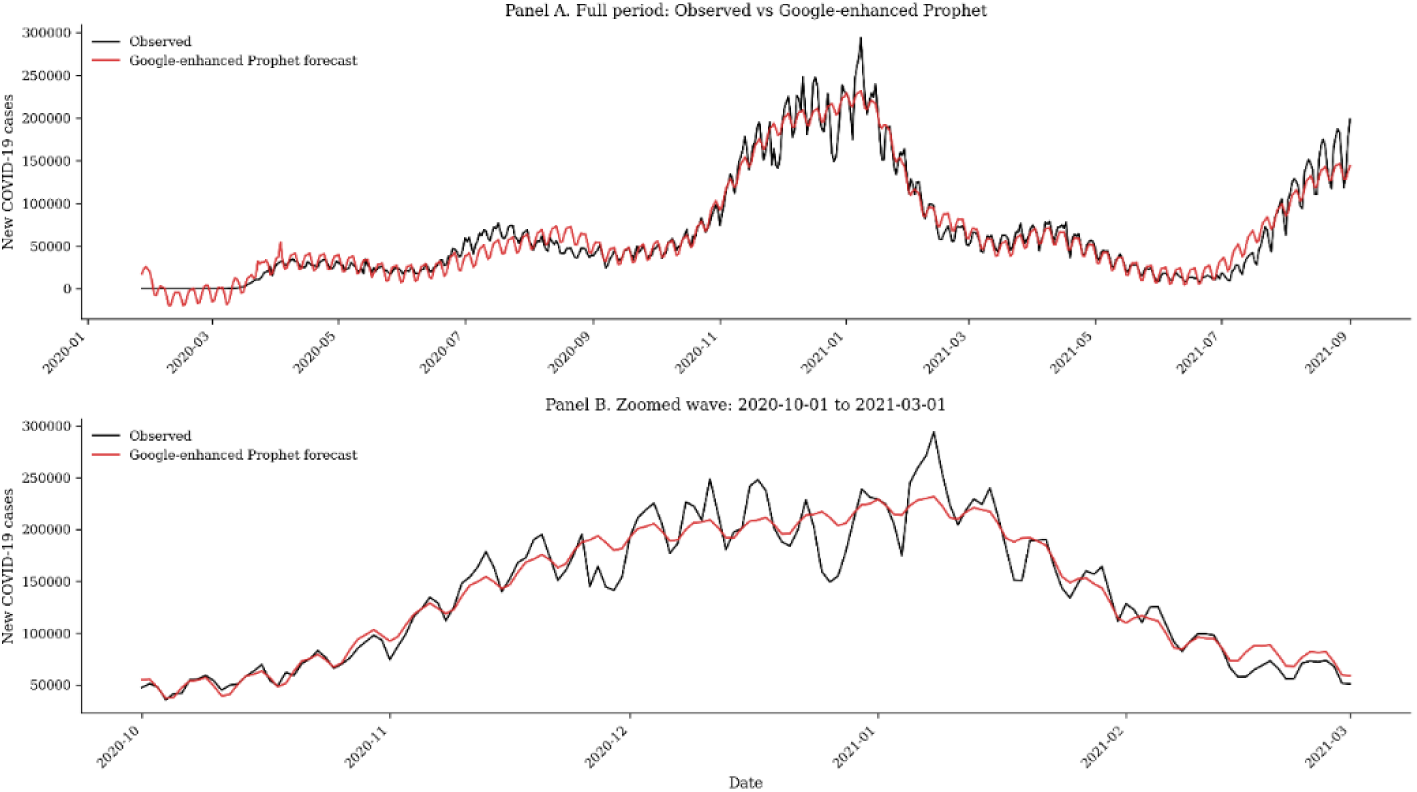
Observed vs Google forecast

**Figure 5:**
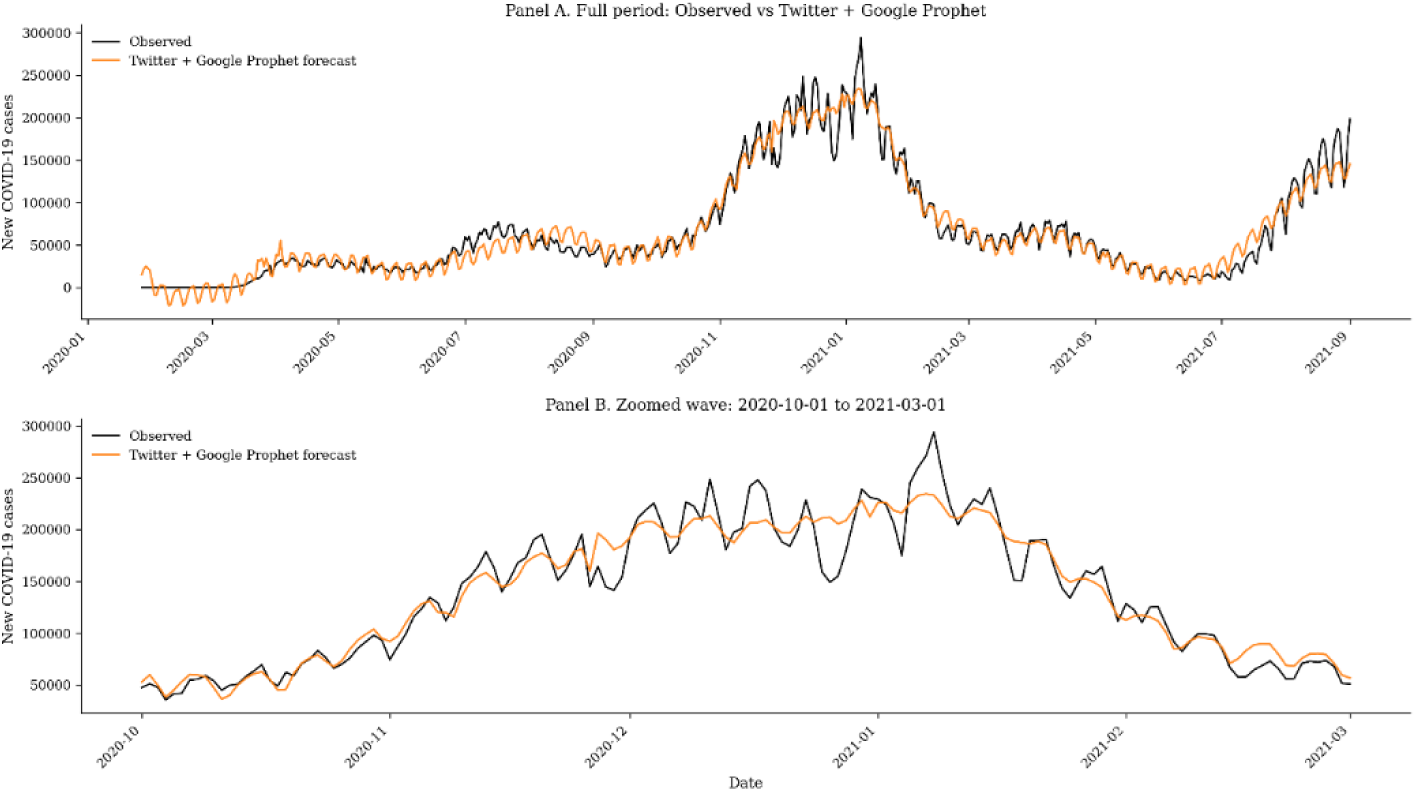
Observed vs Twitter + Google forecast

Twitter-enhanced model more accurately captured both the timing and magnitude of epidemic waves. In contrast, the Google-only model tended to lag behind changes in transmission and underestimated the height of several waves. The combined model followed a trajectory close to the Twitter-enhanced model without offering further accuracy benefits.

To further evaluate how each model performed during periods of rapid epidemiological change, Figure 6 presents smoothed model trajectories across the first three major U.S. COVID-19 waves. These focused views allow assessment of short-term alignment between observed case counts and model predictions during periods when forecasting is typically most challenging.

**Figure 6:**
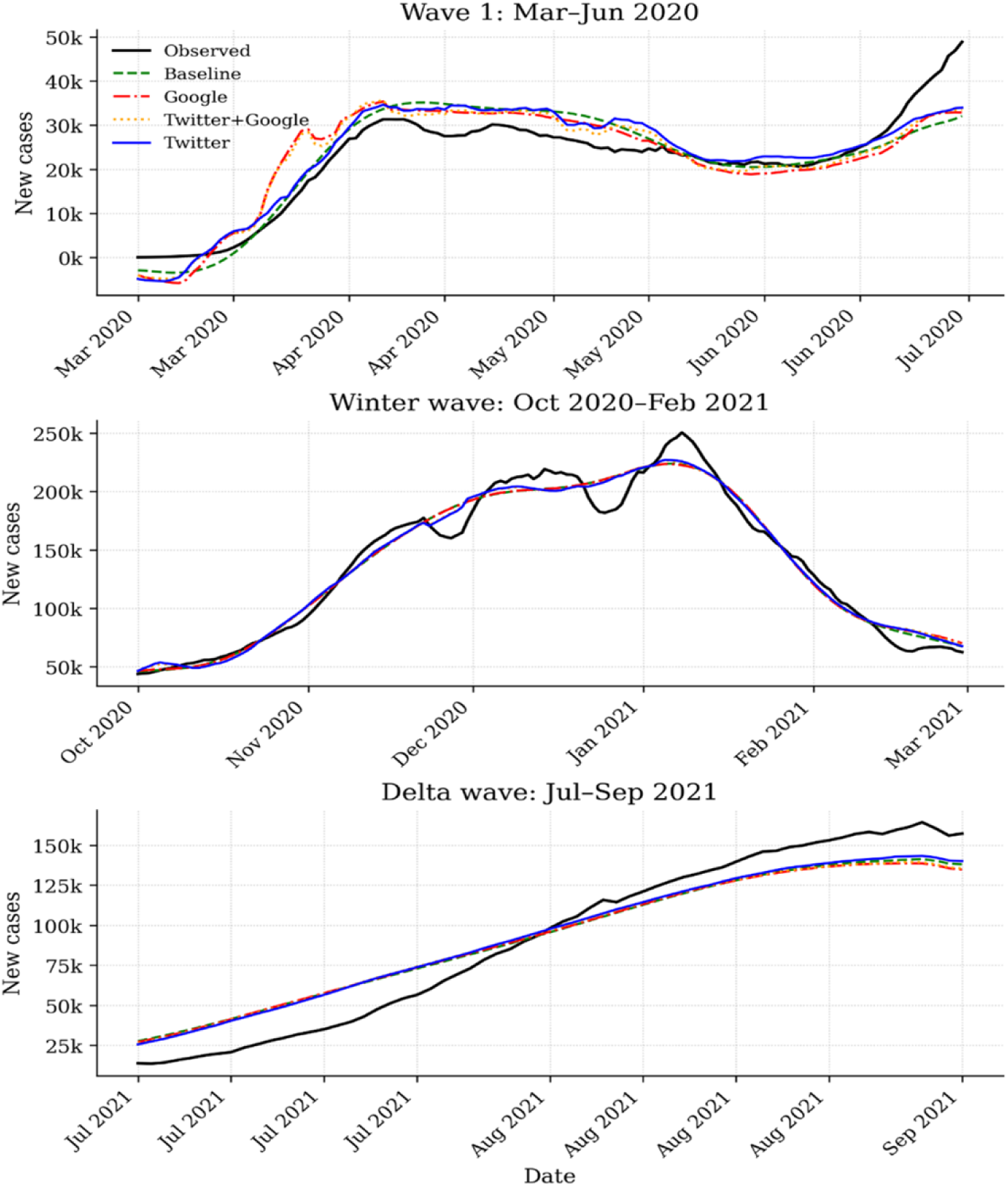
Smoothed model trajectories across US COVID-19 waves

Across all three waves, the Twitter-enhanced model consistently tracked the onset and peak of transmission more closely than the baseline or Google-only models. During the spring and winter waves, Twitter signals enabled faster identification of rising case trends and reduced peak underestimation. The Google-only model frequently exhibited delayed responsiveness and tended to smooth over rapid increases in transmission, resulting in lower peak estimates and a slower decline following each wave. The combined Twitter + Google model largely mirrored the Twitter-only model, suggesting that Google search trends provided limited incremental benefit once emotion-based Twitter regressors were included.

Notably, during the Delta wave (characterized by a steady, sustained rise in incidence) all advanced models performed similarly, though the Twitter-enhanced model remained slightly closer to observed values throughout the period. These results reinforce that social-media-derived regressors, particularly real-time emotion classifications, improve responsiveness and peak approximation during volatile outbreak conditions.

#### Forecast Degradation Across Horizons

The relationship between forecast error and horizon length is displayed in Figure 7. The baseline model showed rapid error growth as the horizon extended beyond approximately ten days. The Twitter-enhanced model exhibited substantially slower degradation in accuracy across the thirty-day horizon. The Google-enhanced and combined models demonstrated intermediate performance, improving accuracy at short horizons but losing stability as the forecast extended farther into the future.

**Figure 7:**
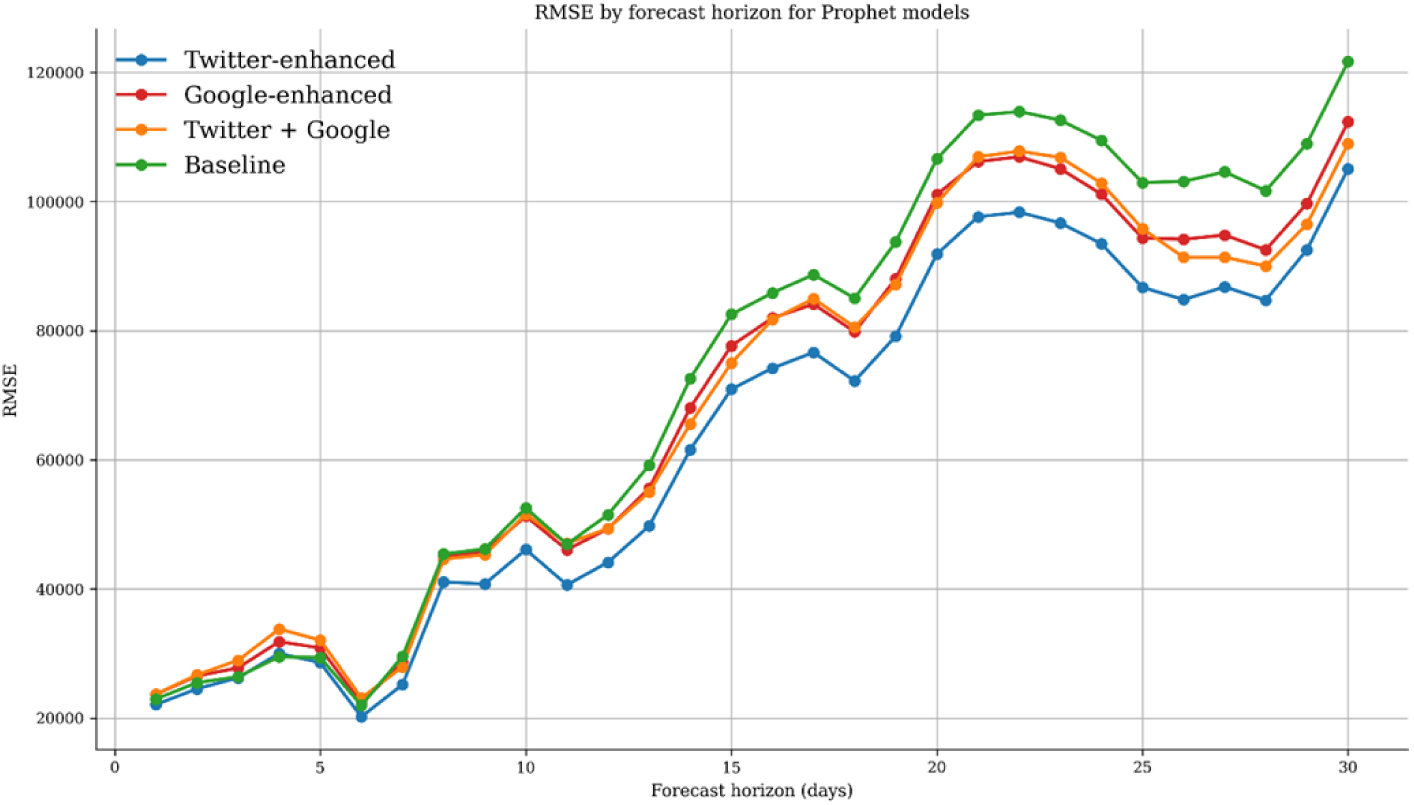
Errors by forecast horizon

### SARIMAX Robustness Test

To assess whether the contribution of digital signals remained consistent across different modeling frameworks, we conducted a robustness test using the SARIMAX models with the same sets of regressors evaluated in the Prophet analysis. Each model was estimated using historical COVID-19 case data and then used to generate 30-day rolling forecasts. These new models’ performances are displayed in table 5.

Overall, the SARIMAX results followed the same broad pattern observed for Prophet. The baseline model, which relied only on historical case counts, produced the highest forecasting error. Incorporating Twitter emotion signals led to modest but consistent improvements, reducing RMSE by about 3% and MAE by 3.2% relative to the baseline. The Google-only model did not improve accuracy and instead slightly degraded performance, showing higher RMSE and MAE than the historical-only model. When Twitter and Google regressors were combined, performance remained close to the Twitter-only model but did not offer additional gains.

These results reinforce that Twitter-derived behavioral signals provide incremental forecasting value beyond historical case trends alone, whereas Google symptom-search indicators were less reliable predictors in this setting. Although the magnitude of improvement was smaller for SARIMAX compared with Prophet, the direction and consistency of effects across models support the robustness of the main findings.

## DISCUSSION

Twitter variables provided the clearest and most consistent improvements in this study. Three categories of tweets, namely Neutral, NoEmotion, and VeryPositive, emerged as the strongest predictors (Neutral tweets correspond to messages that express content with minimal affect but still convey evaluative or informational tone, whereas NoEmotion tweets are classified as lacking detectable emotional expression altogether, often reflecting purely factual reporting, automated updates, or link sharing without subjective framing). This pattern contrasts with some of the early infoveillance literature, which focused heavily on emotional reactions such as fear, anger, or anxiety (11, 29, 42, 43). Several recent studies have noted that neutral or informational posts often rise when people seek practical updates, policy guidance, or local conditions, which can correspond to real epidemiological activity rather than sentiment-driven spikes (12, 44, 45). Our findings strengthen this view by showing that emotion-light content may track meaningful behavioral shifts more reliably than overtly emotional discourse.

Focusing on neutral or emotion-light content may also offer an additional practical advantage for digital epidemiology. Highly emotional social media posts have been shown to disproportionately amplify misinformation and sensationalized narratives, particularly during public health crises (46–48). By emphasizing signals derived from Neutral and NoEmotion tweets, forecasting models may be less exposed to volatility driven by disinformation dynamics and short-lived attention spikes that are weakly related to underlying transmission. Although this study does not directly measure misinformation exposure, the stronger and more stable performance of emotion-light Twitter signals suggests that they may provide a comparatively cleaner behavioral signal for epidemic forecasting (44).

Google Trends indicators showed weaker forecasting value. Although symptoms like cough and anosmia have demonstrated epidemiological relevance in many contexts (20–22, 41), Google search activity is strongly shaped by news reporting, testing policy, diagnostic awareness, and other external forces that introduce noise into predictive models (19, 24, 49). This likely explains why Google regressors did not improve forecasts, especially when combined with Twitter signals. The limited and unstable contribution of Google symptom searches represents an important negative finding, reinforcing concerns raised in prior work about the reliability of search-based indicators for epidemic forecasting. Similar challenges have been described in comparative evaluations of search data during COVID-19 and influenza outbreaks (18, 25, 50).

A notable result is that the improvements associated with Twitter signals were robust across two distinct modeling approaches. Although SARIMAX models achieved lower absolute forecast errors than Prophet, the relative performance gains from Twitter-based regressors were larger and more pronounced in Prophet. Prophet, which adapts well to nonlinear trends and structural changes, showed clearer gains when Twitter regressors were included, whereas SARIMAX, which relies on a more rigid autoregressive structure, exhibited smaller but still consistent improvements. Importantly, the direction and ranking of digital signal effects were consistent across both frameworks, supporting the robustness of the findings. This consistency is important because digital epidemiology studies often rely on a single forecasting model, making it difficult to disentangle model-specific effects from genuinely predictive signals (8, 34, 51–53). By aligning results across model classes, the present study offers stronger evidence that the observed improvements stem from the information embedded in Twitter activity rather than from quirks of any one algorithm.

The visual comparison of epidemic waves suggests the high predictive value of Twitter. While the twitter-enhanced model did not eliminate forecast error during rapid surges, it reacted more responsively to shifts in transmission than the baseline model. This performance aligns with recent work showing that online behavioral data often captures public awareness and behavioral adjustments in the days preceding major epidemiological peaks (54–57).

## LIMITATIONS

The analysis was performed using national COVID-19 case counts, which obscures local variation in transmission. National-level aggregation was chosen to reduce reporting noise and to align with the resolution of the digital indicators, which are most stable at broader spatial scales. However, previous work has shown that the predictive value of digital indicators often changes across spatial scales because online behavior can reflect local concerns and information environments rather than national trends (58). Future work should evaluate whether the advantages of Twitter indicators observed here persist at state or county levels.

There are also limitations associated with the digital datasets themselves. The Twitter data relied on automated emotion classifications, and although these models perform reasonably well, they may misclassify ambiguous or context-dependent expressions. Studies have noted persistent challenges in sentiment and emotion detection when applied to large-scale public health surveillance (59). Google Trends data face their own constraints. They are influenced by various factors which introduce noise that weakens the stability of symptom-search predictors, a pattern observed in other evaluations of Google-based epidemic forecasting tools (60).

The modeling choices used in this study present additional limitations. Prophet and SARIMAX were selected to represent two common forecasting paradigms, yet they do not capture the full range of available approaches. More flexible architectures such as long short-term memory networks or hybrid machine-learning frameworks may identify nonlinear interactions between digital indicators and case dynamics that classical models overlook. Prior research has shown that the performance of digital predictors can vary considerably across model classes, highlighting the value of multimodel comparisons (61).

## CONCLUSION

Across both Prophet and SARIMAX models, Twitter emotion indicators consistently improved predictions, while Google symptom searches showed limited benefit. The consistency of these results across two different modeling approaches suggests that certain forms of online behavior, particularly neutral and informational tweets, may capture early shifts in public awareness that correspond to changes in transmission.

The value of Twitter signals was evident during several epidemic waves and across different forecasting model classes. These findings point to the potential of social media like Twitter as complementary inputs to established forecasting systems. At the same time, the modest and unstable contributions of Google search data highlight the need for caution when using search-based indicators, especially in settings where public attention fluctuates independently of disease activity.

As digital platforms continue to evolve, there is a need for ongoing assessment of how online behavior reflects real epidemiological patterns. Future work should examine how these signals perform at local levels, how they interact with other data sources, and how they can be incorporated into operational forecasting pipelines. The results of this study show that digital traces can add meaningful value, but their usefulness depends on the type of platform, the nature of the signal, and the modeling framework used.

## Data Availability

All datasets used in this study are publicly available: the COVID-19 case data is from the U.S. Centers for Disease Control and Prevention (CDC) (https://covid.cdc.gov/covid-data-tracker), Twitter emotion variables from the Open ICPSR annotated COVID-19 Twitter dataset (https://www.openicpsr.org/openicpsr/project/120321/version/V8/viewjsessionid=4700CD2F12241FE2426BF32C8D257F83?path=/openicpsr/120321/fcr:versions/V8/Twitter-COVID-dataset---Jan-2021), and Google Trends symptom indices are from the COVID-19 Google Symptoms Search Dataset (https://github.com/GoogleCloudPlatform/covid-19-open-data/blob/main/docs/table-search-trends.md)

## ACKNOWLEDGEMENTS

The authors thank the organizations that make COVID-19 surveillance and digital trace data publicly available.

## AUTHORS’ CONTRIBUTIONS

TG led the study design, conducted the data analysis, and drafted the initial manuscript. IB contributed to writing, interpretation of results, and critical review of the manuscript. Both authors read and approved the final version.

## DATA AVAILABILITY

All data used in this study are publicly available. Daily COVID-19 case data were obtained from the U.S. Centers for Disease Control and Prevention COVID Data Tracker. Twitter emotion indicators were sourced from the Open ICPSR annotated COVID-19 Twitter dataset, and symptom search trends were taken from the Google COVID-19 Symptoms Search Dataset. Detailed information on data sources, preprocessing, and reproducibility is provided in the Reproducibility and Code Availability section. No restrictions apply to the access or reuse of the datasets analyzed in this study.

